# Omicron and vaccines: An analysis on the decline in COVID-19 mortality

**DOI:** 10.1101/2022.05.20.22275396

**Authors:** Blas J. Larrauri, Alejandro Malbrán, José A. Larrauri

## Abstract

The SARS-CoV-2 virus emerged in December 2019 infecting more than 430 million people worldwide and causing almost 6 million deaths until February 2022. Rapid vaccination efforts during this period coincided with a reduction in the mortality rate of the virus. A new genetic COVID-19 variant named Omicron appeared and widely spread by the end of 2021, after which the COVID-19 mortality rate showed a marked, albeit temporary, decline. The potential relationship between vaccines and omicron infection on the mortality rate of COVID-19 is analyzed in this article using online data from public sources from countries with relatively high incidence of infection. Mortality and incidence rates were compared before and after Omicron became the prevalent source of COVID-19 cases, as well as the effect of vaccination during these periods. Infection rates were higher during Omicron than in the pre-Omicron period (4.16% vs. 2%, respectively), whereas mortality rates showed the opposite trend both in deaths over population (0.021% vs. 0.171%) and deaths over positive cases (0.27% vs. 1.07%, respectively). The results suggest that vaccines, while significantly reducing mortality, did not prevent Omicron infection; and that during the Omicron period mortality decreased by a low aggressiveness of this variant.

**Key Messages:**

* Vaccines do not appear to have prevented Omicron infection.
* Infection rates were higher during the Omicron period than before it.
* Mortality rates were lower during the Omicron period than before it.
* Vaccines reduced mortality rates during both pre- and Omicron periods.
* The sharp decrease in mortality rates during the Omicron period seems to be due to the low virulence of Omicron strains rather than vaccine efficacy.

## Introduction

A novel human pathogen, the Severe Acute Respiratory Syndrome Coronavirus 2 (SARS-CoV-2) virus, emerged in Wuhan (China) in December 2019 causing the Coronavirus disease 2019 (Zhu et al., 2019; Li et al., 2020; World Health Organization, 2020). The virus quickly spread globally and led to a pandemic, resulting in more than 430 million diagnosed cases and close to 6 million deaths until Feb 2022.

At the beginning of the pandemic the COVID infection did not have a specific treatment, and all efforts were thus focused on the development a vaccine. The first available vaccines received an emergency regulatory approval in late 2020 (Lamb, 2021), after which prompt vaccination strategies were implemented. More than 40 million people worldwide were vaccinated with at least one dose by January 2021 (around 0.5% of world population; Adejumo & Adejumo, 2021), a number that increased to more than 4 billion by November 2021, with 54% with one dose and 42% with two doses (Our World in Data, 2022). Vaccines showed high efficacy in preventing severe COVID-19 infection and mortality (Knoll and Wonodi, 2021; Polack et. al., 2020).

Around November 26 2021 a new COVID-19 variant was identified, named Omicron (World Health Organization, 2021). This COVID-19 mutation was highly transmissible, even in fully vaccinated individuals and people who had received a booster dose (Espenhain et al., 2021; CDC COVID-19 Response Team, 2021). The Omicron variant has shown attenuated replication both in in-vitro (human lung cells; Gupta, 2022; Shuai et al., 2022) and in-vivo studies (mice models; Shuai et al., 2022; Diamond et al.; 2022) when compared to that of SARS-CoV-2 wildtype, as well as an attenuated pathogenicity (Bentley et al., 2021; McMahan et al., 2022). Omicron infection spread globally and became the most prevalent variant of COVID-19 in the world, representing more than 90% of all circulating variants in February 2022 (Our World in Data, 2022).

Simultaneously, COVID-19 infections by the end of 2021 started exhibiting different traits than those seen up to that point, particularly higher rates of infection and lower mortality. The aim of the present study is to analyze the available world-wide COVID data to assess the potential relationship between vaccines and omicron infection on the mortality rate of COVID-19.

## Methods

Data on the Coronavirus pandemic were obtained from the website Our World in Data (https://ourworldindata.org/), from the period spanning from January 22 2020 to February 28 2022. Gathered records included date, location, cases, deaths, and vaccination status.

Incidence of infection (accumulated cases over total population) was assessed on February 28 2022 in order to allocate data from all locations into 4 groups: Group 1 (infection rate lower than 5%), Group 2 (between 5 and 15%), Group 3 (between 15 and 30%), and Group 4 (higher than 30%). Data from groups 1 and 2 were discarded for further analyses since low incidence rates may be indicative of an incomplete description of the situation in the region.

The onset date of the Omicron period was established by determining the time point in which the percentage of cases corresponding to the Omicron variant was significantly larger (> 60%) than the rest of COVID-19 variants. COVID-19 data were then clustered before and after Omicron onset, and arranged by the percentage of population that had been fully vaccinated (i.e., 2 doses) by that date. Pearson correlation coefficients were used to analyze the relationship between vaccination rate and mortality rates pre- and after Omicron onset.

## Results

Two hundred and five locations were included in the initial analysis. When sorting by incidence of infection (see Methods section), 90 countries were included in Group 1, 52 in Group 2, 39 in Group 3, and 24 in Group 4. Groups 3 and 4 (*n*=63) included countries in North and Central America (12), South America (3), Europe (37), Asia (8), Africa (1) and Oceania (2), accounting for a combined population of 1 billion people.

The percentage of COVID-19 variants as a function of time is shown in Figure 1. Delta was the most prevalent variant on December 31 2021 (65% of all cases), being replaced by Omicron at the following time point (January 15 2022, 74%). Interpolation of these values yielded January 9 2022 as the onset date of the Omicron period, when more than 60% of COVID-19 cases corresponded to this latter variant.

**Figure 1:**
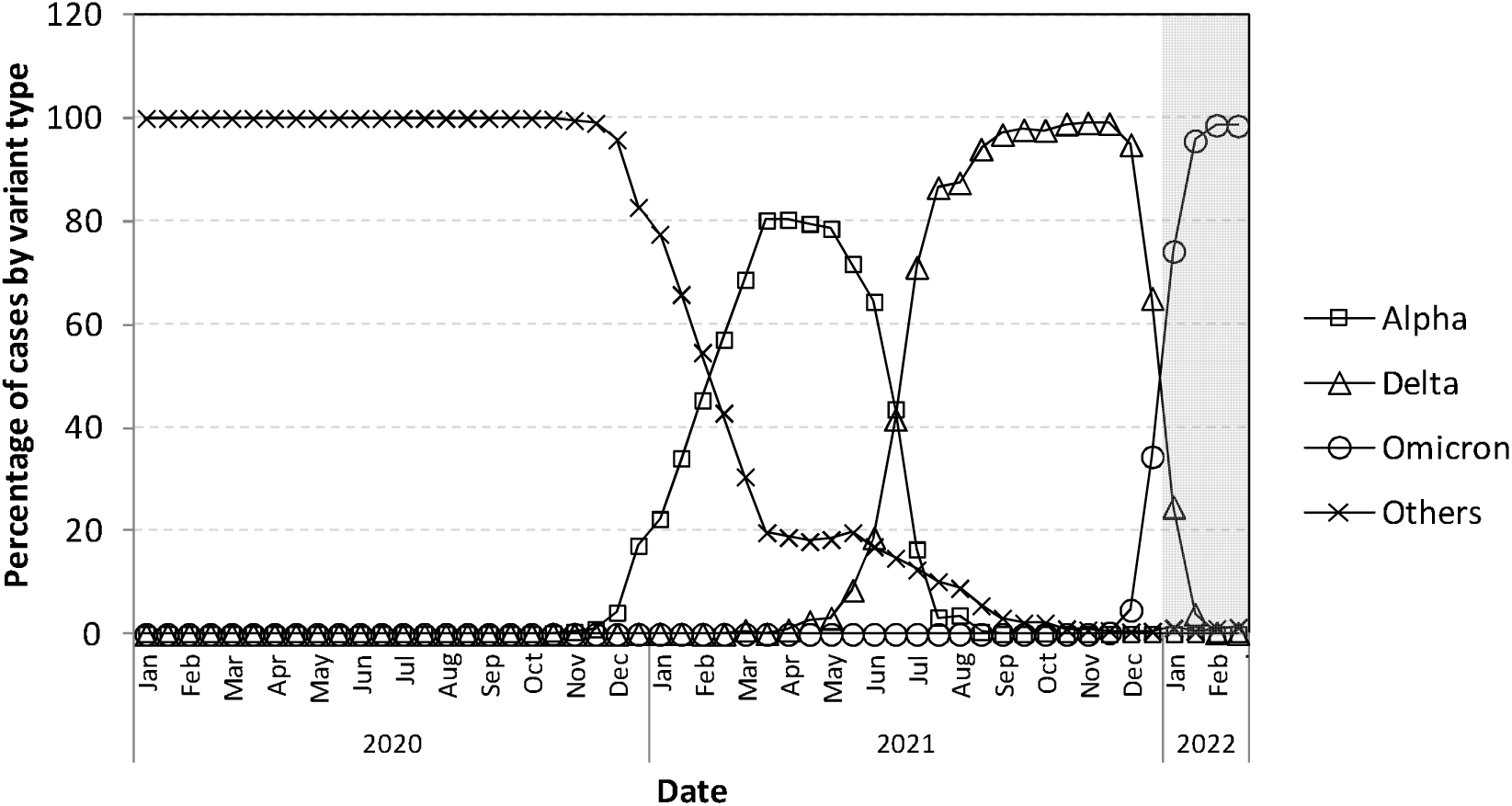
Percentage of COVID-19 cases corresponding to different strains over time. The highlighted area in the figure indicates the period in which Omicron is the prevalent variant (starting on January 9 2022).

The number of biweekly COVID-19 cases per population as a function of time is shown in Figure 2 (open diamonds, left axis). During the pre-Omicron period, biweekly new cases were below 2% of the total population, a rate that spiked during the Omicron stage (4.16%). This increase in COVID-19 cases was reflected in the overall infection rate (accumulated cases over population): 16.76% during the pre-Omicron period (January 22 2020 to January 9 2022, 718 days), and 10.68% during the Omicron phase (January 9 2022 to February 28 2022, 50 days).

**Figure 2:**
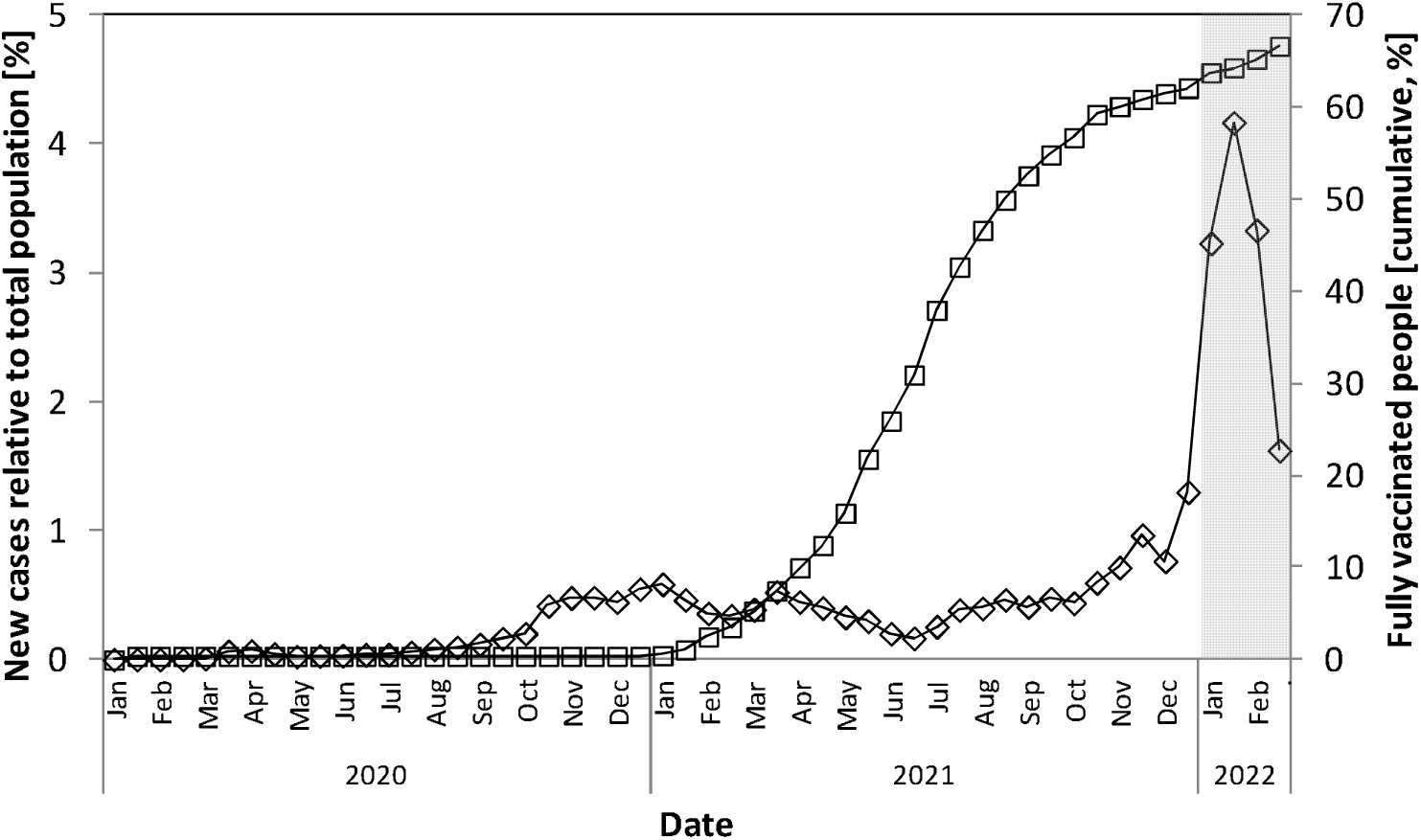
Percentage of new COVID-19 cases (relative to overall population; open diamonds, left axis) and cumulative vaccination rate (open squares, right axis) over time. The highlighted area in the figure corresponds to the Omicron period.

The percentage of fully vaccinated people (shown in Figure 2 -open squares, right axis-) reached around 60% around November 15 2021, and increased slightly to 66.6% by the end of February 2022. Despite the high ratio of people already having received two doses of the vaccine in this 3.5-month period, the onset of the Omicron variant -and the peak of new cases relative to the population-was observed during this stage.

Mortality rate over population showed a steep decline after Omicron onset (pre-: 0.171% vs. Omicron: 0.021%; data not shown). Similarly, mortality rate over positive cases exhibited a marked drop when comparing those periods: 1.07% pre vs. 0.27% during Omicron (Figure 3).

**Figure 3:**
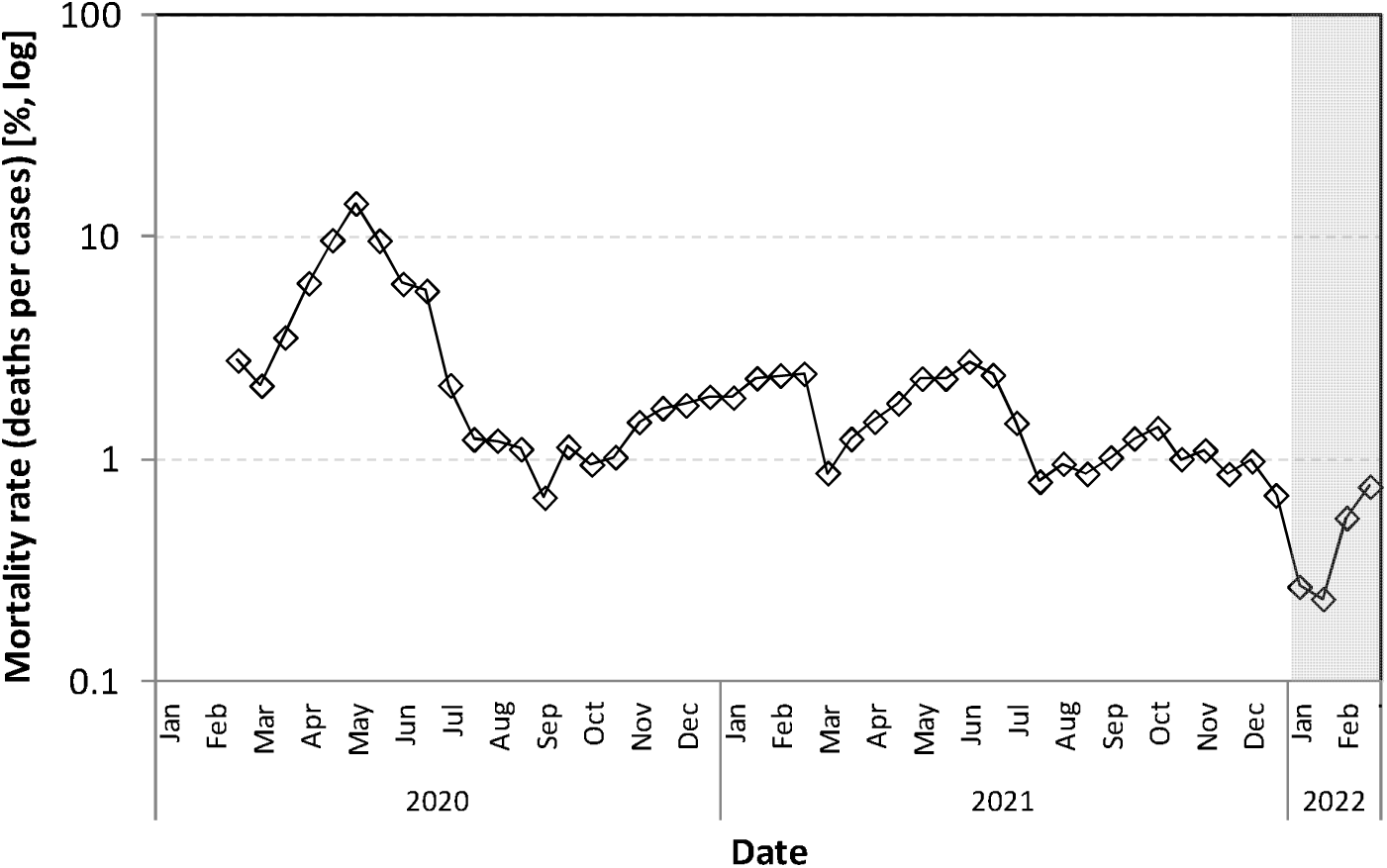
Mortality rate (percentage of deaths relative to the number of positive cases) of COVID-19 over time. The Omicron period is highlighted in the figure.

During the pre-Omicron period, the mortality rate over positive cases was 1.71%, 1.15% and 0.98% for countries with vaccination rates lower than 40%, between 40% and 60%, and higher than 60%, respectively. During Omicron, the mortality rate was 0.46%, 0.44% and 0.22% for those countries. Pearson correlation tests showed a significant negative relationship between vaccination rate and mortality rate pre-Omicron (2 doses before January 9 2022; r(60)=-.275, *p*=.03), as well as during Omicron (r(60)=-.319, *p*=.011), suggesting a decrease in mortality with increasing percentages of vaccinated individuals (see Figure 4). A similar result was obtained when analyzing deaths over total population (data not shown).

**Figure 4:**
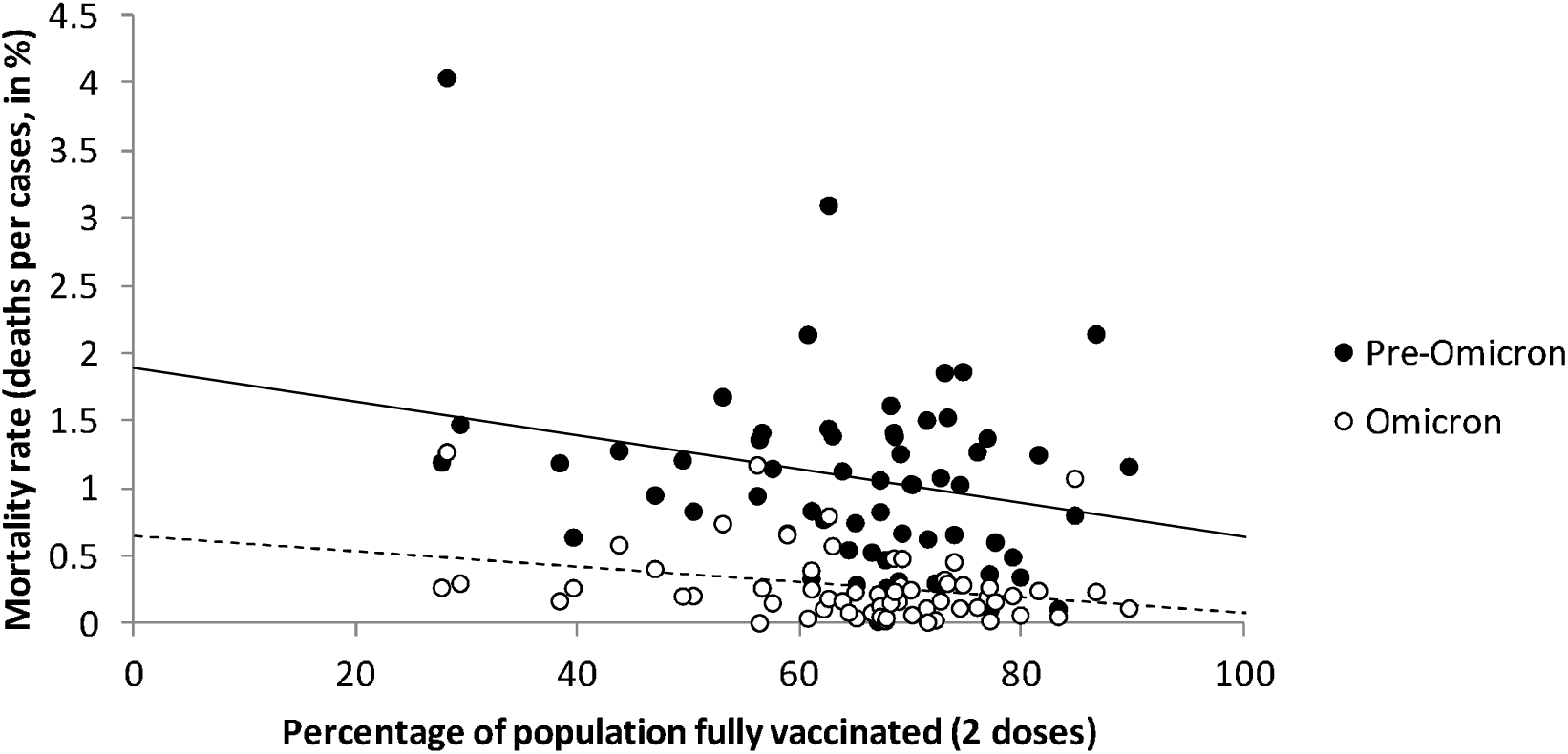
Relationship between vaccination and mortality rate during the pre-Omicron (solid circles; r(60)=-.275, *p*=.03) and Omicron (open circles; r(60)=-.319, *p*=.011) periods. The lines show the corresponding linear regressions.

## Discussion

Mortality rates observed during Omicron were lower than those registered before its onset: 0.021% vs. 0.171% when considering deaths over population, and 0.27% vs. 1.07% over positive cases. Vaccines appear to have an impact in mortality in both pre- and Omicron period, as evidenced by the significant (though small) correlation between vaccination rates and mortality in both of these stages (Figure 4); that is, countries with a larger percentage of people vaccinated exhibited less global and infection mortality.

The sharp decrease in mortality rates observed at the onset of the Omicron period cannot be fully explained by the administration of vaccines: during the pre-Omicron period, average mortality rate ranged from 1.7% (in countries with ∼25% of population fully vaccinated) to 1% (in areas with vaccination above 80%); after Omicron onset mortality those rates decreased to 0.5% and 0.1%, respectively.

As shown in Figure 3, the onset of the Omicron variant coincides with a marked decrease in mortality rate. Similarly, Figure 3 also shows smaller reductions in mortality around March 2021 and July 2021, corresponding to moments in which other variants (Alpha and Delta, respectively) were becoming prevalent (see Figure 1). The reduced mortality rate of Omicron appears to result from an increased contagiousness and lower virus lethality. It has been previously reported that Omicron infection in animal models and in vivo studies exhibited lower mortality than the rest of COVID-19 variants (Diamond et al., 2021; McMahan et al., 2022; Bentley et al., 2021). Interestingly, after the initial drop following Omicron onset, mortality rate appears to recover to its pre-onset values (around 1%) at the end of the Omicron period (Figure 3). This trend also seemed to be present with the Alpha and Delta variants, with an initial decrease in mortality rate at the onset of the variant that faded over time. The extent to which the decline in mortality rate associated to the onset of the Omicron variant persists over time remains uncertain, and warrants further analyses.

The raw data for the analyses in the present report did not include the time of individual vaccination, and therefore it was not possible to determine if the response to vaccines was lost over time. Countries that administered vaccines early may have experienced reduced immune effects during Omicron. Also, distinct vaccines providers and technologies may have different properties (both in infection and mortality), and these factors should be included in future studies analyzing the effectiveness of vaccines in COVID-19. In addition, many countries report all deaths from individuals infected with the SARS-CoV-2 virus as deaths attributable to COVID-19, when the underlying cause of death may differ in patients with severe co-morbidities (Riffe and Acosta, 2021). This practice could result in an artificial increase in COVID-19 mortality rates, especially during Omicron when a substantial part of the population became infected during a short period. Therefore, Omicron mortality may be even lower than that reported.

Examination of available data revealed that a number of countries reported less than 5% of population infected until February 28 2022, with a few indicating less than 1%. In some cases, geographic isolation may explain those low values. However, reduced infection rates may reflect errors in collecting or incomplete accounts, and therefore countries reporting infection rates of less than 15% of their populations were not included in the analyses. Incomplete or reduced infection reports result in larger mortality per positive cases, potentially confounding the results.

Regarding the incidence of COVID-19 infection, the number of cases over total population was slightly higher in during the pre-Omicron period (16.76%) than after Omicron onset (10.68%). However, the former period lasted 718 days whereas the latter was 50 days long, indicating that the infection rate was considerably higher during Omicron (0.213%/day) than before (0.023%/day).

As shown in Figure 2, more than 60% of the population was fully vaccinated at the onset of the Omicron period, when the incidence rate started to rise before reaching a maximum. This peak value was the highest infection rate detected, including those observed during previous variants when vaccination rates were lower. The occurrence of a peak of infection after more than 60% of the population was fully vaccinated suggests that vaccines, despite effectively reducing mortality, were not able to prevent Omicron infection. And notably, if vaccines do not prevent infections of new COVID-19 variants –as appears to have been the case with Omicron-, public policies based on vaccination status, such as health (or vaccination) passes that allow free circulation only to vaccinated individuals, do not seem to be warranted.

## Data Availability

All data was obtained from the website Our World in Data, and all the data is still available on the web

https://ourworldindata.org/

